# Belimumab with rituximab for the treatment of primary membranous nephropathy

**DOI:** 10.64898/2026.08.26.26360913

**Authors:** Sharon A. Chung, Lia Stelzig, Matthew A. Sherman, Wendy Gao, Patricia Tosta, Laura Cooney, Sharon Adler, Nabeel Aslam, Isabelle Ayoub, Andrew S. Bomback, Gaia Coppock, Vimal K. Derebail, Fahmeedah Kamal, Dana V. Rizk, Katherine R. Tuttle, Meryl Waldman, William T. Barry, Patrick H. Nachman

**Affiliations:** Immune Tolerance Network, San Francisco, CA; Rho, Durham, NC; Division of Allergy, Immunology, and Transplantation, National Institute of Allergy and Infectious Diseases, National Institutes of Health, Rockville, MD; Immune Tolerance Network, Ann Arbor, MI; Division of Nephrology, Hypertension, and Transplantation, Department of Internal Medicine, University of California, Los Angeles, Los Angeles, CA; Lundquist Research Institute, Torrance, CA; Department of Nephrology and Hypertension, Mayo Clinic, Jacksonville, FL; Division of Nephrology, Department of Medicine, Ohio State University, Columbus, OH; David Koch Jr. Glomerular Kidney Center, Division of Nephrology, Department of Medicine, Columbia University, New York City, NY; Renal, Electrolyte, and Hypertension Division, Department of Medicine, University of Pennsylvania, Philadelphia, PA; UNC Kidney Center, Division of Nephrology and Hypertension, Department of Medicine, University of North Carolina at Chapel Hill, Chapel Hill, NC; Division of Nephrology, Department of Medicine, Stanford University, Palo Alto, CA; Nephrology Division, Department of Medicine, University of Alabama at Birmingham, Birmingham, AL; Providence Medical Research Center, Providence Inland Northwest Health, Spokane, WA; Division of Nephrology and Kidney Research Institute, Department of Medicine, University of Washington School of Medicine, Seattle, WA; Kidney Diseases Branch, National Institute of Diabetes and Digestive and Kidney Diseases, National Institutes of Health, Bethesda, MD; Division of Nephrology and Hypertension, Department of Medicine, University of Minnesota, Minneapolis, MN

**Author notes:** **Corresponding author:** Sharon A. Chung, MD MAS, Director, Clinical and Translational Medicine Immune Tolerance Network, 535 Mission Street, Suite 2650 San Francisco, CA 94105.

**Keywords:** primary membranous nephropathy, belimumab, rituximab, B cell depletion, anti-PLA2R, clinical trial

## Abstract

**Introduction:** B cell depletion with rituximab leads to complete or partial remission (CR/PR) in only ∼60% of patients with primary membranous nephropathy (PMN). Adding belimumab to rituximab may result in greater depletion of memory B cells, limit the re-emergence of autoreactive B cells, and improve clinical responses.

**Methods:** REBOOT Part A (NCT03949855) is a single arm, open-label, pharmacokinetic study where all participants had proteinuria ≥ 4g/day and detectable serum anti-phospholipase A2 receptor (anti-PLA2R) antibodies. Participants received belimumab 200 mg subcutaneously weekly for 52 weeks and rituximab 1000 mg intravenously at weeks 4 and 6. Assessments included belimumab exposure at week 4 and CR/PR at week 104.

**Results:** Seventeen participants started belimumab. Belimumab exposure was not significantly reduced in those with high (>8 g/day) proteinuria at week 4. Among all treated participants, 59% (10/17) achieved CR/PR at week 104, while in per protocol analyses, 91% (10/11) achieved CR/PR at week 104. All participants in per protocol analyses had normal serum albumin and undetectable serum anti-PLA2R by week 104. Circulating memory B cells increased before rituximab and were depleted by rituximab. B cell re-constitution occurred after week 52 with primarily naïve and transitional B cells. Belimumab with rituximab was well-tolerated, with one participant discontinuing belimumab due to infection.

**Conclusion:** In this study, a high proportion of participants receiving belimumab with rituximab achieved CR/PR. Thus, a multi-targeted approach to B cell depletion may improve immunologic and clinical outcomes in PMN and is being studied in a larger, randomized, placebo-controlled clinical trial.

## Introduction

Primary membranous nephropathy (PMN), an important cause of nephrotic syndrome in adults, results from *in situ* immune complex formation in the subepithelial zone of the glomerular basement membrane causing podocyte injury. Approximately 70% of patients with PMN produce autoantibodies targeting the phospholipase A_2_ receptor (PLA2R), which is expressed by podocytes.^1^

Treatments for patients with PMN with moderate or severe nephrotic syndrome include immunosuppressive agents and non-immunosuppressive anti-proteinuric therapy (e.g., renin angiotensin aldosterone system [RAAS] inhibition). Current treatment guidelines recommend using rituximab (RTX), corticosteroids with cyclophosphamide, or a calcineurin inhibitor^2^ for patients with persistent proteinuria > 3.5 g/day despite non-immunosuppressive anti-proteinuric therapy. With RTX monotherapy, ∼60% of patients attain a complete or partial remission (CR/PR) by 24 months.^3^

Incomplete remission after RTX treatment may be related to incomplete depletion of memory B cells and an increase in B cell activating factor (BAFF) after RTX treatment, which facilitates the survival of autoreactive B cells.^4–6^ Belimumab (BEL), a monoclonal antibody directed against BAFF, is approved for the treatment of systemic lupus erythematosus (SLE). Previous studies suggest that BEL increases the frequency of circulating memory B cells where they may be more susceptible to RTX-mediated depletion. Administering BEL after RTX delays B cell reconstitution, with preferential effects on both naïve and potentially autoreactive B cells.^7–11^ BEL also reduces anti-PLA2R autoantibodies and proteinuria in patients with PMN.^12^ Therefore, initiating BEL prior to RTX may increase B cell depletion. Continuing BEL during and after RTX may limit the re-emergence of naïve and memory B cells, including autoreactive B cells, and improve clinical outcomes in PMN.

REBOOT (“Belimumab and Rituximab Compared to Rituximab Alone for the Treatment of Primary Membranous Nephropathy”, NCT03949855) is a two-part, prospective, phase II, multicenter clinical trial of BEL and RTX in adults with anti-PLA2R positive PMN. Part A is a single-arm, open-label study to assess how BEL exposure is affected by proteinuria, where all participants received BEL and RTX and were followed for clinical outcomes. Part B, which is underway, is a double-blind, randomized, placebo-controlled effectiveness study assessing the effect of adding BEL to RTX in patients with PMN. The BEL exposure analysis as well as the effect of BEL and RTX on clinical outcomes and serologic markers of PMN through week 104 from REBOOT Part A are presented here.

## Methods

REBOOT Part A is an open-label study designed to compare BEL exposure between participants with “low” (≥ 4 to < 8 g/day) and “high” proteinuria (≥ 8 g/day) at study entry. REBOOT is conducted by the Immune Tolerance Network and funded by the Division of Allergy, Immunology, and Transplantation at the National Institute of Allergy and Infectious Diseases. GSK plc provided BEL for the study and had no role in the study design, conduct, data analysis, or drafting of the manuscript.

### Study participants

All participants provided written informed consent, and the institutional review board at each of the 10 sites across the United States approved the study. Participants in Part A were between the ages of 18-75 years and had detectable serum anti-PLA2R antibodies at screening. Participants had to have one of the following: (1) PMN confirmed by kidney biopsy within the past 5 years, (2) PMN confirmed by a kidney biopsy within the past 7 years that is relapsing following CR/PR, (3) nephrotic syndrome with an estimated glomerular filtration rate (eGFR) > 60 mL/min/1.73m^2^ and no history of immunosuppressant treatment for nephrotic syndrome, or (4) nephrotic syndrome and a contraindication to kidney biopsy. While on maximally tolerated RAAS blockade, participants had to have an eGFR ≥ 30 mL/min/1.73 m^2^; systolic blood pressure ≤ 140 mmHg and diastolic blood pressure ≤ 90 mmHg; and either proteinuria ≥ 4 and < 8 g/day for at least the previous 3 months or ≥ 8 g/day at study entry.

The main exclusion criteria were secondary MN, RTX use within the previous 12 months, RTX use greater than 12 months ago with undetectable CD19 B cell counts at study entry or a lack of clinical response to RTX, cyclophosphamide use within the past 3 months, and other immunosuppressant use within the past 30 days. A full listing of the entry criteria is provided in the Supplemental Materials Section S1.

### Study treatment and clinical assessments

All Part A participants received 200 mg of BEL subcutaneously weekly for 52 doses and 1000 mg of RTX intravenously at weeks 4 and 6. BEL 200 mg administered weekly was the approved subcutaneous dose for the treatment of SLE at the start of the study. Following completion of the 52-week treatment phase, participants were followed for another 52 weeks off all immunosuppressant treatment.

The first 4 BEL injections were performed at the study site under the supervision of a healthcare provider. Trough serum BEL levels were obtained weekly following the first 4 doses of BEL and before the administration of RTX. Serum BEL trough levels were assessed using a validated electrochemiluminescence immunoassay (LabCorp Early Development Laboratories, Chantilly, VA).

Clinical assessments including physical exams, adverse events (AEs), and laboratory studies occurred every 4 weeks during the first 52 weeks of the study and quarterly from weeks 52-104. Proteinuria quantified by 24-hour urine collection and serum anti-PLA2R antibodies were assessed quarterly from study enrollment until week 104. The Kidney Disease Quality of Life-36 instrument (KDQOL-36, www.rand.org/health-care/surveys_tools/kdqol.html)^13^ was assessed quarterly through week 52 and semiannually through week 104.

Protocol-specified reasons for discontinuing belimumab included worsening kidney function, worsening proteinuria, and relapse (definitions provided in Supplemental Materials Section S2). Participants who discontinued BEL entered safety monitoring and had AEs, serum chemistry, immunoglobulin levels, and CD19 counts assessed quarterly until week 104. Proteinuria and anti-PLA2R titers were not assessed in safety monitoring.

Clinical laboratory assessments were performed by a central laboratory (LabConnect, Johnson City, TN), except for anti-PLA2R antibody assessments, which were performed by ARUP Laboratories (Salt Lake City, UT) using a semi-quantitative cell-based indirect fluorescent antibody assay. A titer of <1:10 was considered negative.

The primary clinical outcome was the proportion of participants in CR/PR at week 104. CR was defined as proteinuria ≤ 0.3 g/day with serum albumin ≥ 3.5 g/dL, and PR was defined as a 50% or greater decrease in proteinuria compared to baseline and proteinuria < 3.5 g/day, as previously defined in the MENTOR study.^3^ The definitions of CR and PR were updated mid-study, as described in Supplemental Materials Section S2. Secondary clinical outcomes included the level of proteinuria and proportion of participants with negative serum anti-PLA2R at weeks 52 and 104.

### Mechanistic assessments

Serum and peripheral blood mononuclear cells (PBMCs) were collected at study entry and at least quarterly until week 104. Mechanistic assessments included flow cytometry of B cell subset frequencies.

PBMCs were isolated by density gradient centrifugation and stored in liquid nitrogen for batched analysis. For analysis, PBMCs were thawed and stained with the antibody panel shown in Supplemental Table S2. Data was acquired on a CyTEK Aurora Spectral Cytometer and unmixed using SpectrFlo (CyTEK). Hierarchical gating for B cell subsets was conducted in FlowJo V10 (Supplemental Figure S1). B cell subsets assessed include naïve (CD19+ CD27-IgD+ CD38-CD24-), transitional (CD19+ CD27-IgD+ CD38+ CD24+), switched memory (CD19+ CD27+ IgD-CD38-), unswitched memory (CD19+ CD27+ IgD-), double negative (CD19+ CD27-IgD-), and antibody-secreting cells (CD19+ CD27+ IgD-CD38hi).

Data was analyzed using GraphPad Prism 10. Relative B cell subset frequencies were converted to absolute cell number per microliter of blood using absolute CD19+ cell counts from clinical labs. B cell subset analysis was limited to samples with at least 100 B cells captured by flow cytometry. Not all participants had PBMCs samples and absolute B cell count data available at all timepoints. Therefore, the number of data points across timepoints varied from five to 12. Figures present data for participants who were not in safety monitoring at that timepoint.

### Statistical analysis

#### i. Belimumab exposure

REBOOT Part A investigated whether BEL dosing should be increased in patients with significant proteinuria, as BEL was detected in the urine of patients with proteinuria and the urinary excretion of BEL was associated with 24 hour proteinuria levels.^12^ Because BEL pharmacokinetics exhibits dose proportionality^14^, a decrease of 50% in trough concentrations in those with high compared to low proteinuria would justify doubling the dose to 400 mg/week. Twenty participants, with 10 in each proteinuria group, was estimated to provide 90% power to detect a 50% reduction in BEL trough concentrations, assuming a one-sided alpha = 0.1 and coefficient of variation (CV) of 63.3%, based on a multi-dose study of BEL in participants with SLE.^14^

BEL exposure analyses included all treated participants who received BEL through week 4 and had available trough samples. A repeated measures mixed model was applied to BEL trough levels on the natural log scale, with covariates for baseline proteinuria (low versus high), study week, and their interaction. The estimated ratio of the geometric means between the high and low proteinuria groups is reported with a 95% confidence interval (CI). An additional mixed model evaluated baseline proteinuria as a continuous variable, and an exploratory break point analysis considered all possible proteinuria cut points in the analyzed participants.

#### ii. Clinical and patient reported outcomes

Part A was not powered to conduct hypothesis tests of clinical outcomes. Descriptive statistics are reported for the treated group and the per-protocol groups at week 52 (PP52) and week 104 (PP104). The treated group includes all participants who initiated treatment with BEL. The per-protocol group includes participants who had no major deviations that impacted clinical outcome assessments through the specified study week, had the study visit at the specified week, received both RTX infusions, and received at least 42 of 52 BEL doses. The proportions of participants in CR/PR are reported at weeks 52 and 104; participants missing remission status assessments were counted as not having achieved CR/PR. Summary statistics are provided for clinical measures including eGFR, serum albumin and the KDQOL-36. All analyses were performed using SAS 9.4.

## Results

### Study enrollment

From February 2020 through May 2022, 26 individuals were screened, 17 initiated BEL, and 12 completed BEL (Figure 1). Five participants discontinued BEL prior to week 52 and entered safety monitoring. The reasons for discontinuation are provided in Figure 1, with additional details provided in Supplemental Materials Section S4. Two of the 5 participants did not receive any RTX, and the other 3 received both doses of RTX. Ten participants were in the PP52 sample, and 11 were in the PP104 sample (Figure 1).

**Figure 1.**
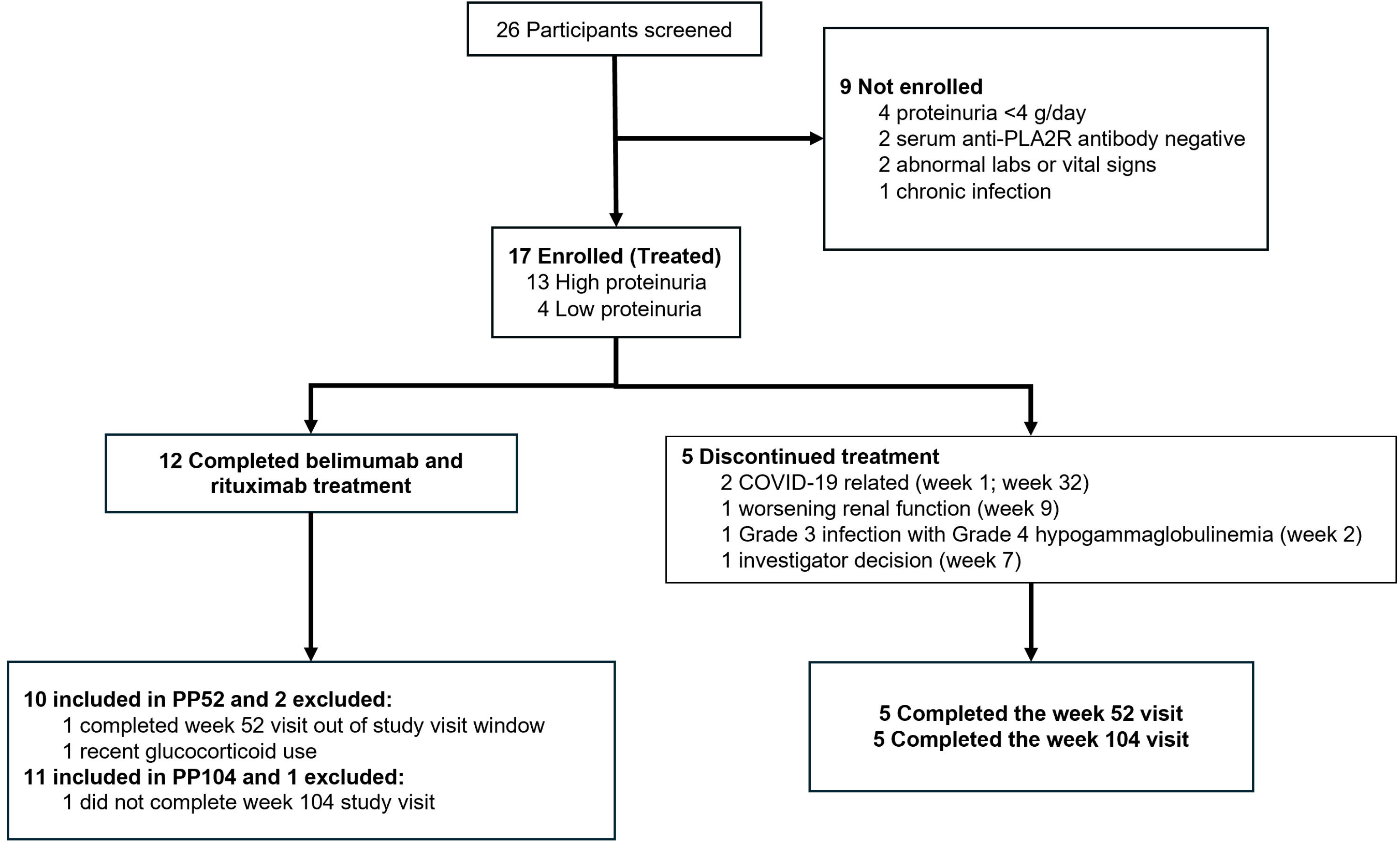
Participant flow diagram for REBOOT Part A.

Participant demographics are presented in Table 1. Most participants were male (82%) and White (82%). The median age at study entry was 58 years (interquartile range [IQR] 55-62). The median proteinuria at screening was 11.3 g/day (IQR 8.2-16.7).

**Table 1.** Participant demographics and laboratory values at study entry.

|  | **Treated**<br>**Population**<br>**(n=17)** | **BEL Exposure Population**<br>**High Proteinuria**<br>**(n=8)** | **Low Proteinuria**<br>**(n=4)** |
| --- | --- | --- | --- |
| Age (years), median (IQR) | 58 (55-62) | 58.5 (55.5-66.0) | 49.5 (33.5-59.5) |
| Male, n (%) | 14 (82) | 6 (75) | 4 (100) |
| Race, n (%)<sup>1</sup> |  |  |  |
| White | 14 (82) | 6 (75) | 3 (75) |
| Black | 2 (12) | 2 (25) | 0 |
| Asian | 1 (6) | 0 | 1 (25) |
| Native Hawaiian or Other Pacific Islander | 1 (6) | 0 | 0 |
| Ethnicity, n (%) |  |  |  |
| Hispanic or Latino | 2 (12) | 1 (13) | 0 |
| Not Hispanic or Latino | 15 (88) | 7 (88) | 4 (100) |
| BMI (kg/m<sup>2</sup>), median (IQR) | 33.0 (30.9-36.1) | 33.6 (31.7-36.6) | 30.4 (24.4-36.4) |
| Weight (kg), median (IQR) | 103.9 (98.8-109.5) | 104.0 (99.9-109.6) | 91.8 (78.1-111.3) |
| Systolic Blood Pressure (mmHg), median (IQR) | 138 (130-149)<sup>2</sup> | 147 (136-150)<sup>2</sup> | 126 (123-133) |
| Diastolic Blood Pressure (mmHg), median (IQR) | 80 (78-82) | 81 (79-84) | 80 (74-83) |
| Proteinuria (g/24 hr), median (IQR) | 11.3 (8.2-16.7) | 14.7 (9.9-17.5) | 6.0 (5.0-6.5) |
| Urine Protein/Creatinine Ratio (mg/g), median (IQR) | 8272 (4953-9725) | 8593 (6612-11212) | 3914 (2955-4954) |
| Albumin (g/dL), median (IQR) | 2.6 (2.3-2.9) | 2.6 (2.4-2.9) | 3.1 (2.4-3.7) |
| Creatinine (mg/dL), median (IQR) | 1.3 (1.0-1.6) | 1.5 (1.2-1.8) | 0.9 (0.8-1.0) |
| eGFR (mL/min/1.73m<sup>2</sup>), median (IQR) | 61.6 (56.9-76.7) | 61.3 (51.7-72.5) | 102.3 (91.9-103.8) |
| Immunoglobulin G (mg/dL), median (IQR) | 371 (223-466) | 380 (305-456) | 501 (289-790) |
| IgG > 690 (Normal), n (%) | 2 (12) | 0 | 1 (25) |
| IgG 550-690 mg/dL (Grade 1), n (%) | 1 (6) | 0 | 1 (25) |
| IgG 400-549 mg/dL (Grade 2), n (%) | 5 (29) | 4 (50) | 0 |
| IgG 250-399 mg/dL (Grade 3), n (%) | 4 (24) | 3 (37) | 1 (25) |
| IgG <250 mg /dL (Grade 4), n (%) | 5 (29) | 1 (13) | 1 (25) |
| Total Cholesterol (mg/dL), median (IQR) | 219 (202-309) | 273 (213-324) | 197 (171-235) |
| LDL Cholesterol (mg/dL), median (IQR) | 129 (115-154) | 138 (126-164) | 118 (98-138) |
| Anti-PLA2R Titer, n (%) |  |  |  |
| 1:40 | 3 (18) | 2 (25) | 1 (25) |
| 1:80 | 2 (12) | 0 | 1 (25) |
| 1:160 | 4 (24) | 1 (13) | 1 (25) |
| 1:320 | 1 (6) | 1 (13) | 0 |
| 1:640 | 3 (18) | 1 (13) | 0 |
| 1:1280 | 3 (18) | 2 (25) | 1 (25) |
| 1:2560 | 1 (6) | 1 (13) | 0 |
| Months From Last Renal Biopsy to Treatment Initiation, median (IQR) | 3.7 (2.7-8.7) | 5.8 (2.8-8.7) | 3.1 (2.5-7.9) |
| Primary MN Diagnosis Year, n (%) |  |  |  |
| 1998-2002 | 4 (24) | 2 (25) | 1 (25) |
| 2003-2007 | 1 (6) | 0 | 0 |
| 2008-2012 | 1 (6) | 0 | 1 (25) |
| 2013-2017 | 0 | 0 | 0 |
| 2018-2022 | 11 (64) | 6 (75) | 2 (50) |

<sup>1</sup>More than one race may be selected by each participant.
<sup>2</sup>All participants fulfilled eligibility criterion of SBP <140 at screening, with median SBP of 129 mm Hg (IQR 120-137 mmHg) at screening; the study protocol did not reassess eligibility at subsequent study visits.

### Belimumab exposure analysis

Due to slow enrollment into Part A, an *ad hoc* interim analysis of BEL exposure was conducted after 12 participants had provided a week 4 BEL trough sample. Eight were in the high proteinuria group and 4 in the low proteinuria group (Table 1).

Table 2 provides geometric mean BEL trough levels after the first 4 doses. The ratio of the mean BEL trough levels after the fourth dose between the high vs. low proteinuria groups was estimated by the mixed model to be 0.79 (95% CI 0.51-1.21). When analyzing proteinuria as a continuous variable, each 1 g/day increase in baseline proteinuria was estimated to result in a 2.0% decrease in trough levels (p=0.27, Figure 2). An exploratory breakpoint analysis did not identify an alternative cut point in baseline proteinuria that significantly differentiated BEL trough levels.

**Figure 2.**
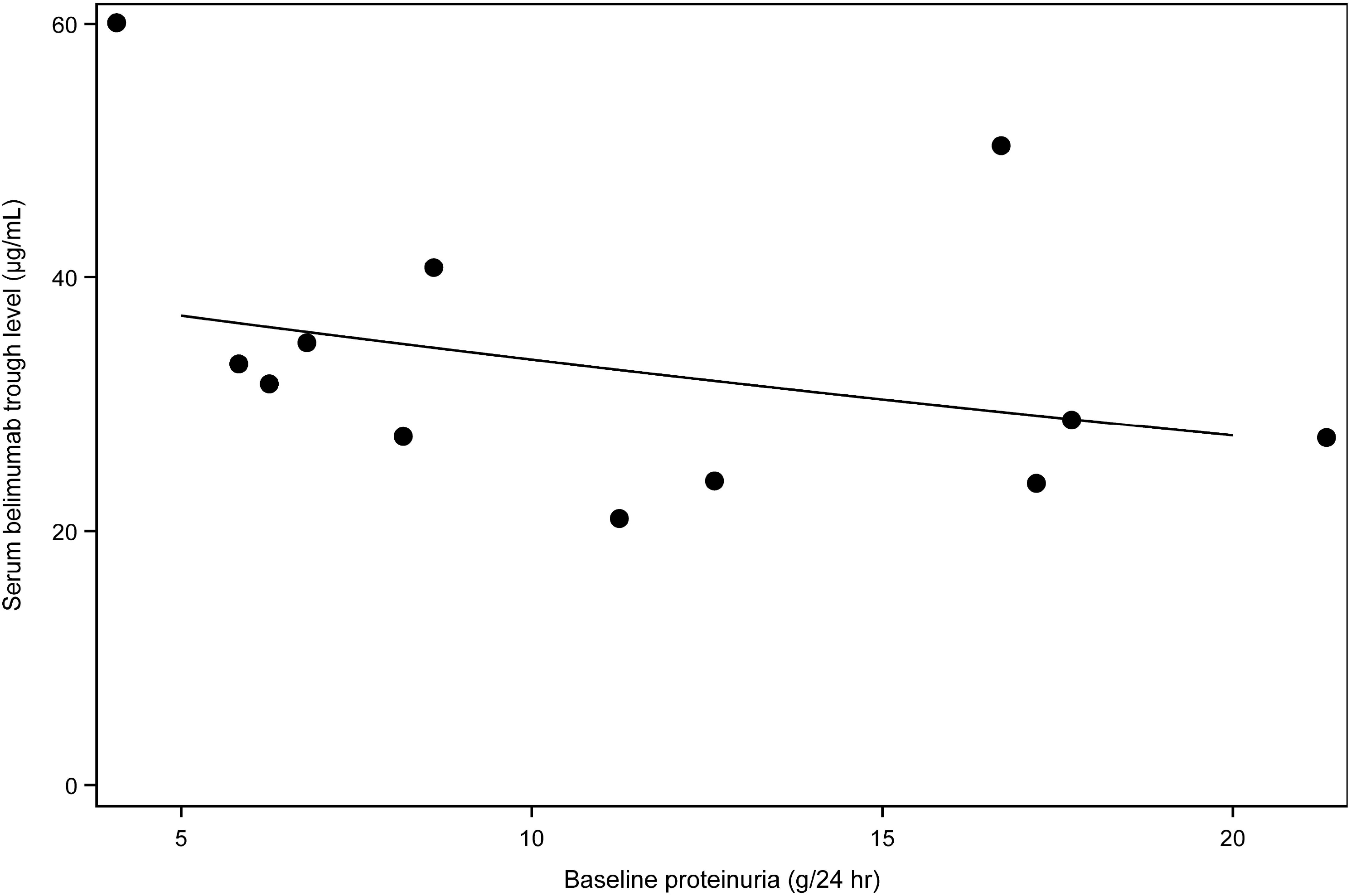
Serum belimumab trough levels after the fourth dose by baseline proteinuria levels. The line presents the predicted belimumab trough level based on a repeated measures mixed model with baseline proteinuria as a continuous variable.

**Table 2.** Geometric mean (% coefficient of variation) of the belimumab trough levels (µg/mL) by proteinuria group.

| Proteinuria level | Week 1 | Week 2 | Week 3 | Week 4 |
| --- | --- | --- | --- | --- |
| High (n=8) | 18.9<sup>1</sup> (23.1) | 23.8 (26.4) | 27.6 (15.2) | 29.2 (32.8) |
| Low (n=4) | 13.99 (44.1) | 24.90 (24.2) | 34.52 (28.8) | 38.51 (33.7) |

<sup>1</sup>At Week 1, there were 7 assessments in the high proteinuria group, as one outlier trough level measurement was judged to be a technical error and excluded from analysis

Based on the observed trough levels, the 95% prediction interval was 0.56 to 1.02 for the ratio of geometric means at the target sample size of 20 participants. The predictive probability of observing a ratio of geometric means of 0.5 or smaller at the target sample size was less than 0.01. Thus, these results did not support a higher dose of belimumab for participants with “high” baseline proteinuria.

### Clinical and patient reported outcomes

Participants treated with BEL and RTX had a rapid decline of serum anti-PLA2R antibodies (Figure 3d). Seven of 12 (58%) participants and 11 of 13 (85%) participants had undetectable anti-PLA2R antibodies at week 12 and 24, respectively. Ten out of 12 (83%) participants had undetectable serum anti-PLA2R antibodies at week 52. All 11 (100%) assessed participants had undetectable serum anti-PLA2R antibodies at week 104. One participant who achieved an undetectable anti-PLA2R did not maintain undetectable anti-PLA2R levels throughout study follow-up. For this participant, the anti-PLA2R antibody became undetectable at week 24, and was detectable at week 36 through the end of follow-up at week 91.

**Figure 3.**
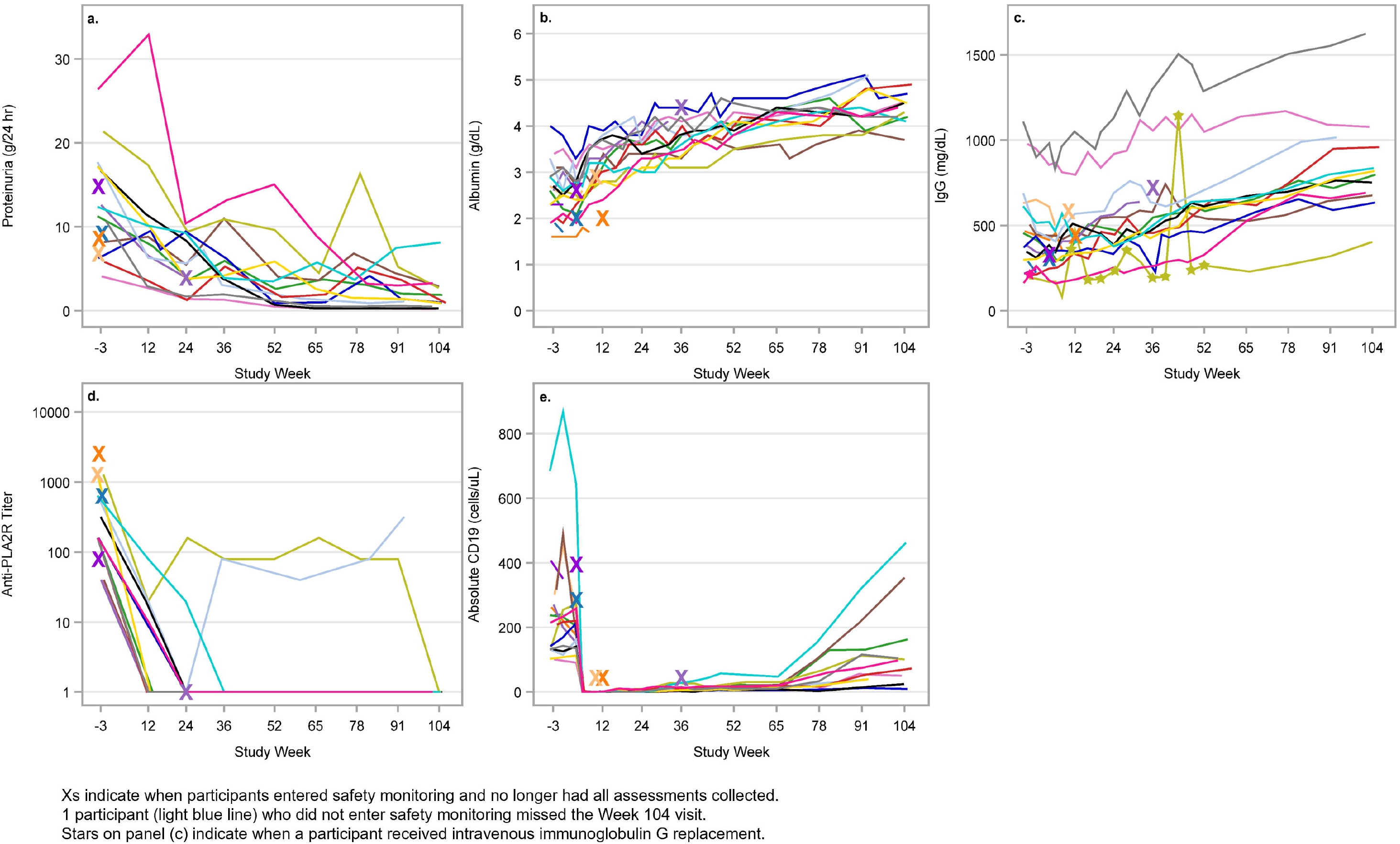
Clinical outcomes for each participant in REBOOT Part A. Each colored line represents the same participant across graphs. (a) 24-hour proteinuria. (b) Serum albumin. (c) Serum immunoglobulin G levels. (d) Serum anti-PLA2R titers, which indicate a ratio (i.e., a value of 80 represents a titer of 1:80). (e) Absolute CD19 B cell counts.

Proteinuria, as assessed by 24-hour urine collection, gradually decreased for all study participants after initiation of study treatment (Figure 3a). The improvement in proteinuria lagged behind the improvement in anti-PLA2R antibody levels. Proteinuria continued to decrease for some participants after stopping study treatment at week 52.

Serum albumin improved after initiation of study treatment (Figure 3b). All participants who completed study treatment normalized their serum albumin level by week 52, and it remained in the normal range even after stopping study treatment through week 104 (Table 3). Serum IgG levels also improved after initiation of study treatment (Figure 3c and Table 3).

**Table 3.**
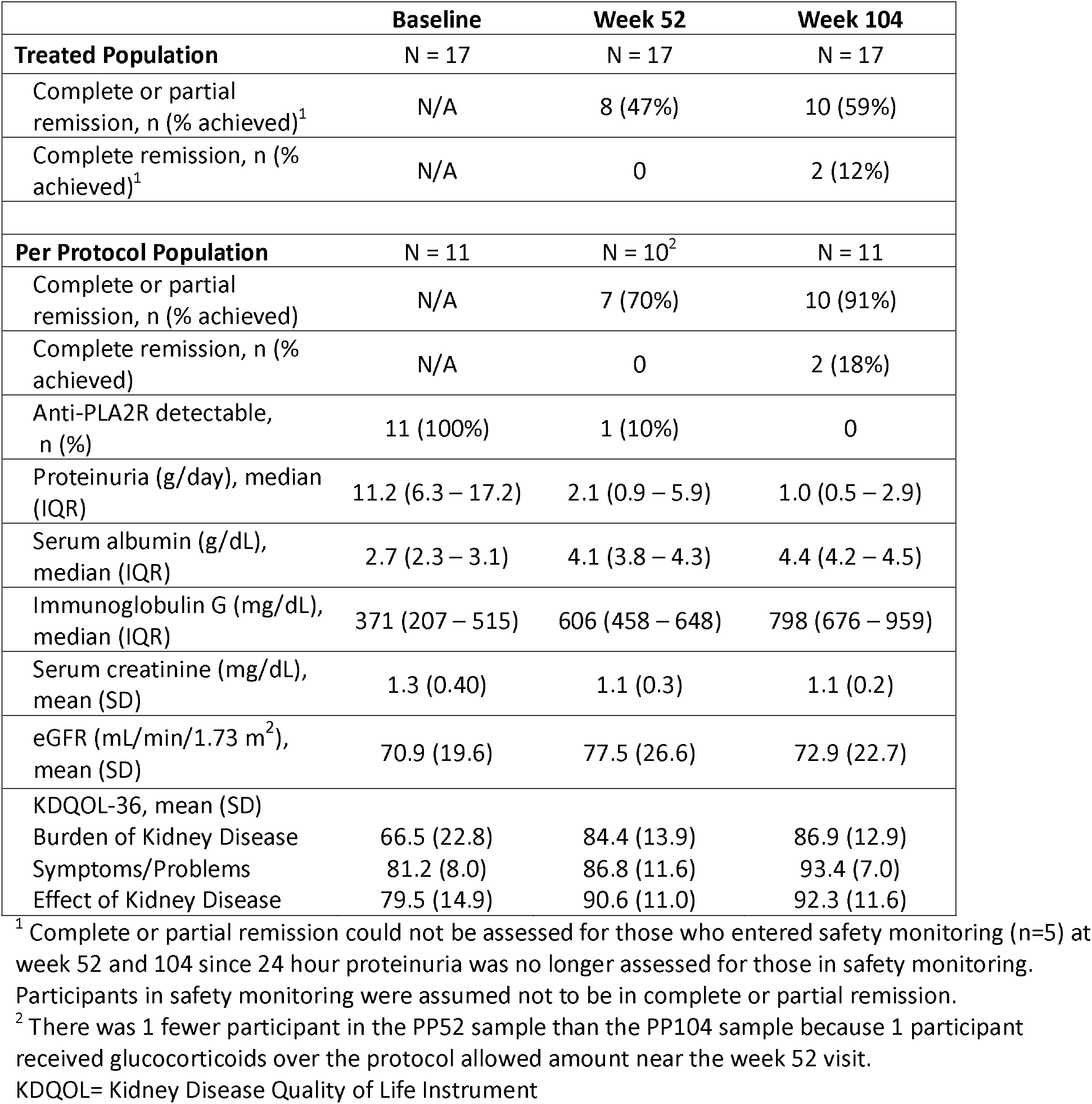
Clinical outcomes for REBOOT Part A participants.

|  | **Baseline** | **Week 52** | **Week 104** |
| --- | --- | --- | --- |
| **Treated Population** | N = 17 | N = 17 | N = 17 |
| Complete or partial remission, n (% achieved)<sup>1</sup> | N/A | 8 (47%) | 10 (59%) |
| Complete remission, n (% achieved)<sup>1</sup> | N/A | 0 | 2 (12%) |
| **Per Protocol Population** | N = 11 | N = 10<sup>2</sup> | N = 11 |
| Complete or partial remission, n (% achieved) | N/A | 7 (70%) | 10 (91%) |
| Complete remission, n (% achieved) | N/A | 0 | 2 (18%) |
| Anti-PLA2R detectable, n (%) | 11 (100%) | 1 (10%) | 0 |
| Proteinuria (g/day), median (IQR) | 11.2 (6.3 – 17.2) | 2.1 (0.9 – 5.9) | 1.0 (0.5 – 2.9) |
| Serum albumin (g/dL), median (IQR) | 2.7 (2.3 – 3.1) | 4.1 (3.8 – 4.3) | 4.4 (4.2 – 4.5) |
| Immunoglobulin G (mg/dL), median (IQR) | 371 (207 – 515) | 606 (458 – 648) | 798 (676 – 959) |
| Serum creatinine (mg/dL), mean (SD) | 1.3 (0.40) | 1.1 (0.3) | 1.1 (0.2) |
| eGFR (mL/min/1.73 m<sup>2</sup>), mean (SD) | 70.9 (19.6) | 77.5 (26.6) | 72.9 (22.7) |
| KDQOL-36, mean (SD) |  |  |  |
| Burden of Kidney Disease | 66.5 (22.8) | 84.4 (13.9) | 86.9 (12.9) |
| Symptoms/Problems | 81.2 (8.0) | 86.8 (11.6) | 93.4 (7.0) |
| Effect of Kidney Disease | 79.5 (14.9) | 90.6 (11.0) | 92.3 (11.6) |

<sup>1</sup> Complete or partial remission could not be assessed for those who entered safety monitoring (n=5) at week 52 and 104 since 24 hour proteinuria was no longer assessed for those in safety monitoring. Participants in safety monitoring were assumed not to be in complete or partial remission.
<sup>2</sup> There was 1 fewer participant in the PP52 sample than the PP104 sample because 1 participant received glucocorticoids over the protocol allowed amount near the week 52 visit.
KDQOL= Kidney Disease Quality of Life Instrument

Circulating CD19 B cells became undetectable after initiation of RTX (Figure 3e) and were not detected again until around week 65.

When assessing all treated participants, 8 of 17 (47%) achieved CR/PR at week 52 and 10 of 17 (59%) achieved CR/PR at week 104. This analysis included those in safety monitoring who were not included in the per protocol groups and assumed those in safety monitoring did not achieve CR/PR. CR and PR could not be formally assessed since urine studies were not obtained for participants in safety monitoring. Renal function (assessed by serum creatinine and eGFR) remained stable for all participants during the study period.

In per-protocol analyses, 7 of 10 (70%) of the PP52 participants achieved CR/PR by week 52. Among the 11 participants in the PP104 group, 10 (91%) had achieved CR/PR by week 104, of which 2 (18%) achieved CR (Table 3). One participant who did not achieve CR/PR at week 104 achieved PR at week 52 with proteinuria of 3.48 g/day (baseline 12.4 g/day), but then proteinuria increased to 8.11 g/day by week 104. This participant’s anti-PLA2R titers were undetectable, serum albumin had normalized, and serum creatinine was similar to baseline at week 104. A kidney biopsy obtained prior to week 104 demonstrated PLA2R-positive staining by immunofluorescence with intramembranous and subepithelial deposits as well as glomerulomegaly and peri-hilar focal segmental glomerulosclerosis.

To assess the effect of study treatment on the participant’s quality of life, participants completed the KDQOL-36 during the study. Quality of life scores improved in all three domains (burden of kidney disease, symptoms/problems, and effect of kidney disease) from baseline to week 104. Most of the improvement occurred between baseline and week 52 (Table 3). The greatest magnitude of improvement was in the Burden of Kidney Disease domain. In all three domains, the mean score increased by over 10 points from baseline to week 104, which is considered a large effect.^15^

### Safety and tolerability

Through week 104, 96 AEs occurred after treatment initiation, of which 13 events were grade 3 and 7 events were grade 4 (Table 4). No grade 5 AEs occurred. Thirty-eight AEs were considered possibly related or related to BEL. Of the 20 infectious AEs that occurred in 9 participants, 1 was a grade 3 infection while all others were grade 1 or 2. Eight cases of non-serious COVID-19 infections occurred among 6 participants. Supplemental Table S4 provides the categories of AEs that occurred in > 10% of participants.

**Table 4.** Treatment emergent adverse events in REBOOT Part A.

|  | **Participants**<br>**(n=17)** | **Events** |
| --- | --- | --- |
| Total Serious Adverse Events (SAEs), n (%) | 2 (12) | 4 |
| SAEs Related to Belimumab | 1 (6) | 2 |
| SAEs Related to Rituximab | 0 | 0 |
| Total Adverse Events (AEs) | 17 | 96 |
| AEs Related to Belimumab, n (%) | 13 (76) | 38 (40) |
| AEs Related to Rituximab, n (%) | 10 (59) | 27 (28) |
| AEs by Severity, n (%) |  |  |
| Grade 1 | 6 (35) | 7 (7) |
| Grade 2 | 15 (88) | 69 (72) |
| Grade 3 [1] | 8 (47) | 13 (14) |
| Grade 4 [2] | 6 (35) | 7 (7) |
| Grade 5 | 0 | 0 |
| Infection, n (%) | 9 (53) | 20 (21) |

[1] Grade 3 adverse events included: Blood creatinine increased, Bursitis infective, Glomerular filtration rate decreased (4), Hyperkalemia, Hypogammaglobulinemia (3), Hyponatremia, Neutrophil count decreased, Renal impairment
[2] Grade 4 adverse events included: Hypogammaglobulinemia (3), Neutrophil count decreased (3), Retinal detachment

The extent of hypogammaglobulinemia at baseline is shown in Table 1. To meet AE criteria, hypogammaglobulinemia had to increase in grade from baseline. There were 8 hypogammaglobulinemia AEs in 8 participants, and the maximum grade was 3 or higher in 6 participants. Immunoglobulin replacement therapy for hypogammaglobulinemia was started at the discretion of the treating physician. Three participants with grade 4 hypogammaglobulinemia received intravenous immunoglobulin but none of the instances was in response to infection: one received a single dose during screening, one received intermittent doses from study days 76-364, and one received a single dose on day 61 after entering safety monitoring on day 6 (Figure 3c). During the study, IgG improved in all participants who had hypogammaglobulinemia at baseline and remained in the study past week 8.

The only cancer reported, excluding non-melanoma skin cancer, was one case of prostate cancer at week 101. There were no serious injection or hypersensitivity reactions. There were four serious AEs (SAEs) among two participants; one participant had grade 2 retinal artery occlusion and grade 4 retinal detachment, and one participant had grade 3 infective bursitis as well as grade 2 pneumonia. The two infection SAEs were deemed to be related to BEL.

The combination of BEL and RTX overall was well-tolerated. Of the 5 participants who discontinued BEL prior to week 52, only one discontinuation was attributed to the potential toxicity of combining B-cell targeting therapies (the grade 3 infection in the setting of grade 4 hypogammaglobulinemia).

### Mechanistic assessments

PBMC samples were analyzed by flow cytometry to assess relative proportions and absolute frequencies of B cell subsets after treatment with BEL and RTX (representative gating in Supplemental Figure S1). Figure 4A shows the percent change from baseline in absolute frequency (# cells/microliter of blood) of six B cell subsets: switched memory, unswitched memory, antibody secreting cells, naïve, transitional, and double negative. Treatment with BEL prior to RTX was associated with an increased frequency of circulating switched memory, unswitched memory, and double negative B cells at week 4. At week 24, after RTX, all B cell subsets were nearly undetectable. Transitional B cells were the first to repopulate, increasing in number beginning around week 52, returning to pre-treatment levels in most participants around week 78, and increasing to above baseline levels by week 91. Naïve B cells began to repopulate around week 91 and by week 104 had returned to approximately half the baseline level for most participants. Memory and double negative B cells remained rare through week 104. Antibody secreting cells were partially depleted beginning at week 4, possibly related to the direct depleting effect of BEL, but the degree of long-term depletion of antibody secreting cells was highly variable.

**Figure 4.**
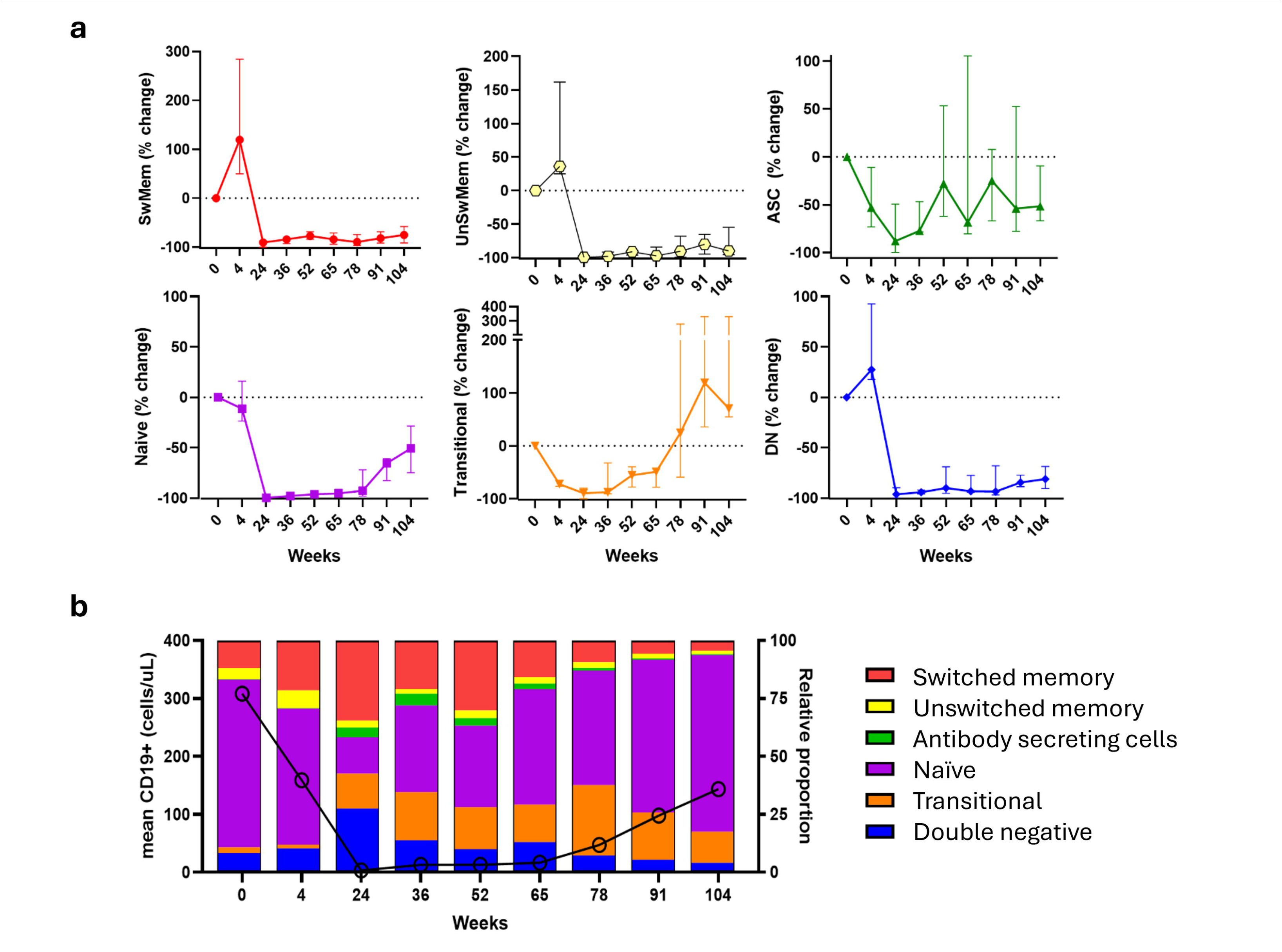
Effects of belimumab and rituximab on B cell subsets. (a) Percent change from baseline in the absolute frequency (number of cells per microliter of blood) of six B cell subsets: SwMem (switched memory), UnSwMem (unswitched memory), ASC (antibody secreting cells), naïve, transitional, and DN (double negative). Representative gating in Supplemental Figure S1. N = 5-12 for each timepoint. Symbols and bars depict median and interquartile range, respectively. Dashed line at zero depicts no change from baseline. (b) Mean absolute CD19+ cell count per microliter of blood (left Y-axis, line plot with hollow circles) and average relative proportion (percent of total B cells) for each of the six B cell subsets (right Y-axis and legend, stacked bars) Color coding in (b) matches (a).

Figure 4B shows the average relative proportion of each of the six B cell subsets in circulation at each timepoint. The average absolute B cell count is overlaid to provide context to relative proportions at timepoints when total counts were low. Comparing week 0 to week 4 demonstrated the increased proportion of circulating memory B cells following treatment with BEL alone, and comparing week 0 to week 104 confirmed the decreased proportion of memory B cells and increased proportion of naïve and transitional B cells. Supplemental Figure S2 shows that the degree of total CD19+ B cell depletion for all participants and the trajectory of reconstitution based on response status.

## Discussion

This study is the first to explore the effect of combining two B cell targeting therapies—BEL and RTX—for the treatment of PMN. Anti-PLA2R autoantibodies were rapidly depleted for almost all participants who received BEL and RTX, and the depletion persisted for at least one year after cessation of immunosuppression. The decrease in autoantibody production was followed by a reduction in proteinuria as well as normalization of serum albumin and IgG levels. Quality of life scores also improved with BEL and RTX. A low rate of serious infections was observed. Overall, these results suggest that utilizing both BEL and RTX may improve clinical outcomes safely in patients with PMN.

Flow cytometry assays supported the underlying study hypotheses—namely, that BEL increases the number of circulating memory B cells that could be depleted by RTX, and continuation of BEL after RTX results in B cell reconstitution of primarily transitional cells, with delayed reconstitution of naïve and memory B cells. In a previous study of RTX in PMN, transitional B cells returned to baseline levels around month 6, whereas transitional B cell recovery was delayed until month 18 in REBOOT Part A. Similarly, naïve B cells reached approximately 50% of their baseline numbers at month 6 after RTX alone, which did not occur until month 24 in REBOOT Part A.^16^ These results are consistent with findings in lupus nephritis.^9,17^ Thus, the addition of BEL before and after RTX in patients with PMN prolongs B cell depletion and drives preferential reconstitution with an immature B cell population.

The demographics of the study population in REBOOT Part A are consistent with those of PMN in general.^18^ At study entry, REBOOT part A participants had moderate or high risk disease^18^ based on proteinuria (median 11.2 g/d) and hypoalbuminemia (median 2.6 g/dL), but had relatively preserved eGFR (median 61.6 ml/min/1.73m^2^). All participants had circulating anti-PLA2R antibodies, and high baseline anti-PLA2R levels are associated with a lower likelihood of spontaneous remission and/or response to RTX.^19–21^ In aggregate, participants had a low likelihood of spontaneous remission based on their degree of proteinuria, hypoalbuminemia, and anti-PLA2R levels. Nevertheless, 91% of per protocol sample participants achieved PR or CR. These results indicate that BEL with RTX can halt anti-PLA2R autoantibody production and subsequently induce PR or CR in patients with moderate to severe PMN.

The observed rate of PR or CR in REBOOT Part A in the per protocol population was 91% at week 104, and all participants in PR at week 104 also had a serum albumin > 3.5 mg/dl. PR with normalized serum albumin has previously been associated with a reduced risk of relapse and improved patient and kidney survival compared to PR with persistent hypoalbuminemia.^22^ This rate of PR or CR in REBOOT Part A is higher than the rate observed in the RTX-treated group from the MENTOR study (60%), which helped establish the role of RTX in the treatment of PMN. However, the rate of CR was lower in REBOOT Part A participants (18%) compared to the rituximab arm in MENTOR (35%). The differences between REBOOT Part A and MENTOR outcomes should be interpreted with caution given the small sample size of REBOOT Part A. However, REBOOT Part A participants at baseline had more severe proteinuria (11.2 g/day vs 8.9 g/day), lower eGFR (68.7 vs 84.9 mL/min/1.73 m^2^), and a higher proportion of participants with circulating anti-PLA2R (100% vs 77%) than RTX-treated MENTOR participants. A significant proportion of MENTOR participants also received a second course of RTX, which was not given in REBOOT Part A.^3^

While nephrotic syndrome is associated with hypogammoglobulinemia, this complication is not well studied in adults with PMN. Almost all (88%) participants in this study had serum IgG levels below the lower limit of normal at baseline, and 13 of 17 participants had baseline levels < 500 mg/dl. This was an important consideration as both BEL and RTX can cause secondary hypogammaglobulinemia. However, serum IgG levels substantially improved during the treatment period and continued to improve after completion of BEL.

REBOOT Part A does have limitations. The sample size is small, which limits the generalizability of the results. The unblinded nature of the study may have affected the assessment of patient reported outcomes but is less likely to impact the primary clinical outcomes that are laboratory-based. The KDQOL-36 is commonly used to assess patient-reported outcomes for patients on dialysis, but it is not validated for patients with PMN. While treatment details before enrollment were not available, the protocol excluded patients with recently treated with immunosuppression, making it unlikely that the observed responses are a residual effect of prior therapy. Finally, as a single arm study, the results of REBOOT Part A cannot be compared with another treatment strategy and are to be confirmed in the ongoing larger, randomized, placebo-controlled REBOOT Part B effectiveness study.

While the primary intent of REBOOT Part A was to assess BEL exposure in individuals with significant proteinuria, the clinical outcomes of this study suggest the potential effectiveness of this treatment strategy. Overall, the results of REBOOT Part A reinforce the concept that a multi-targeted approach to B cell depletion may substantially improve immunologic and clinical outcomes in PMN.

*Funding*: The REBOOT study is conducted by Immune Tolerance Network (ITN), which is supported by the National Institute of Allergy and Infectious Diseases (NIAID) within the National Institutes of Health (NIH) grant UM1AI109565. The study is supported by the NIAID awards UM2AI117870 to the ITN and 75N93022C0000 to Rho, the statistical and clinical coordinating center. Lastly, this research is supported in part by the Intramural Research Program of the National Institute of Diabetes and Digestive and Kidney Diseases (NIDDK) within the NIH. The contributions of the NIH authors are made as part of their official duties as NIH federal employees, are in compliance with agency policy requirements, and are considered Works of the United States Government. However, the findings and conclusions presented in this paper are solely those of the authors and do not necessarily reflect the views of the NIH or the U.S. Department of Health and Human Services.

Belimumab for the study was provided by GSK, plc. GSK was provided the opportunity to review a preliminary version of this publication for factual accuracy. The authors are solely responsible for final content and interpretation.

## Data sharing statement

Data from the REBOOT study presented in this manuscript will be openly available at https://www.itntrialshare.org/ upon publication of the manuscript.

## Acknowledgements

We would like to acknowledge Linna Ding, MD PhD, for her contributions to the development of this study.

## Supplementary Material

Supplementary Material has:

- Section S1: REBOOT entry criteria
- Section S2: Definitions of clinical outcomes

- Supplemental Table S1
- Section S3: Supplemental information for flow cytometry assays

- Supplemental Figure S1
- Supplemental Table S2
- Supplemental Figure S2
- Section S4: Participant discontinuation

- Supplemental Table S3
- Section S5: Treatment-emergent adverse events with incidence ≥ 10%

- Supplemental Table S4)
- STROBE Checklist

Supplementary information is available at the KI Report’s website.

