## Supplemental Materials for "Belimumab with rituximab for the treatment of primary membranous nephropathy"

**Supplemental materials for “Belimumab with rituximab for the treatment of primary membranous nephropathy: initial results of the REBOOT clinical trial”**

**Contents:**

**Section S1:** REBOOT entry criteria

**Section S2:** Definitions of clinical outcomes (Supplemental Table S1)

**Section S3:** Supplemental information for flow cytometry assays (Supplemental Figure S1, Supplemental Table S2, and Supplemental Figure S2)

**Section S4:** Participant discontinuation (Supplemental Table S3)

**Section S5:** Treatment-emergent adverse events with incidence $\geq 10\%$ (Supplemental Table S4)

STROBE checklist

#### Section 1: Entry Criteria from REBOOT Protocol V5.0

##### Inclusion Criteria

Patients *must meet all* of the following criteria to be eligible for this study:

1. Age 18 to 75 years inclusive
2. Diagnosis of one of the following:
  a. Primary membranous nephropathy (MN) confirmed by a kidney biopsy within the past 5 years
  b. Primary MN that is relapsing following a complete remission (CR) or partial remission PR confirmed by a kidney biopsy within the past 7 years
  c. Nephrotic syndrome with eGFR > 60 mL/min/1.73m<sup>2</sup> and no history of immunosuppressant treatment (e.g. glucocorticoids, cyclophosphamide, cyclosporine A, tacrolimus, B-cell depleting agent) for nephrotic syndrome, and without evidence of a secondary cause of nephrotic syndrome
  d. Nephrotic syndrome and a contraindication to kidney biopsy (e.g., anti-coagulation, solitary kidney, body habitus that increases the risk of biopsy, or other contraindication in the opinion of the investigator), and without evidence of a secondary cause of nephrotic syndrome
1. Serum anti-PLA2R positive
2. eGFR ≥ 30 mL/min/1.73m<sup>2</sup> while on maximally tolerated RAS blockade
3. Proteinuria:
  a. ≥ 4 and < 8 g/day that has persisted for at least the previous 3 months while on maximally tolerated RAS blockade. Documentation of persistent proteinuria may be from a 24-hour collection or calculated from a spot urine collection. Or,
  b. ≥ 8 g/day while on maximally tolerated RAS blockade
4. Blood pressure while on maximally tolerated RAS blockade:
  a. Systolic blood pressure ≤ 140 mmHg
  b. Diastolic blood pressure ≤ 90 mmHg
5. SARS-CoV-2 vaccination according to the current Centers for Disease Control and Prevention Advisory Committee on Immunization Practices recommendations. The last SARS-CoV-2 vaccine dose must have been administered at least 14 days prior the initiation of the study drug (Visit 0).

##### Exclusion Criteria

Patients who *meet any* of the following criteria will *not* be eligible for this study:

1. Secondary cause of MN (e.g., SLE, drug, infection, malignancy) suggested by review of the patient's medical history and/or clinical presentation
2. Rituximab use within the previous 12 months
3. Rituximab use > 12 months ago:
  a. With an undetectable CD19 B cell count, or

b. Did not result in a CR or PR with rituximab treatment alone (e.g., without other immunosuppressive or immunomodulatory therapy)
4. Use of anti-B cell therapy other than rituximab within the previous 12 months (or 5 half-lives, whichever is greater)
5. Cyclophosphamide use within the past 3 months
6. Use of other immunosuppressive medications such as cyclosporine or tacrolimus within the past 30 days
7. Use of systemic corticosteroids within the past 30 days
8. Use of any biologic investigational agent (defined as any drug not approved for sale in the country it is used) in the previous 12 months
9. Use of any non-biologic investigational agent in the past 30 days (or 5 half-lives, whichever is greater)
10. Poorly controlled diabetes mellitus defined as hemoglobin A1c (HbA1c) $\geq 9.0\%$
11. Patients with diabetic glomerulopathy on renal biopsy that is:
  a. Greater than Class I diabetic glomerulopathy, or
  b. Class I diabetic glomerulopathy with a history of poor diabetic control (e.g., HbA1c $\geq 9.0\%$) since time of biopsy
12. Unstable kidney function defined as $> 20\%$ decrease in eGFR during the previous 3 months due to primary MN, as determined by the site investigator in consultation with the protocol chair
13. Decrease in proteinuria by 50% or more during the previous 12 months
14. WBC count $< 3.0 \times 10^3/\mu\text{l}$
15. Absolute neutrophil count $< 1.5 \times 10^3/\mu\text{l}$
16. Moderately severe anemia (hemoglobin $< 9 \text{ g/dL}$)
17. History of primary immunodeficiency
18. Serum IgA $< 10 \text{ mg/dL}$
19. Alanine aminotransferase (ALT) or aspartate aminotransferase (AST) $\geq 2\text{x}$ the upper limit of normal (ULN)
20. Positive HIV serology
21. Positive HCV serology, unless treated with anti-viral therapy with achievement of a sustained virologic response (undetectable viral load 24 weeks after cessation of therapy)
22. Evidence of current or prior infection with hepatitis B, as indicated by positive HBsAg or positive HBcAb
23. Positive QuantiFERON – TB Gold test results. PPD tuberculin test may be substituted for QuantiFERON – TB Gold test
24. History of lung disease with FVC $< 70\%$ predicted, DLCO $< 70\%$ predicted, or requiring supplemental oxygen
25. History of malignant neoplasm within the last 5 years except for basal cell or squamous cell carcinoma of the skin treated with local resection only or carcinoma *in*

*situ* of the uterine cervix treated locally and with no evidence of metastatic disease for 3 years

26. Absence of individualized, age-appropriate cancer screening
27. Women of child-bearing potential who are pregnant, nursing, or unwilling to be sexually inactive or use FDA-approved contraception until week 104
28. Acute or chronic infection, including current use of suppressive therapy for chronic infection, hospitalization for treatment of infection in the past 60 days, or parenteral anti-microbial (including anti-bacterial, anti-viral, or anti-fungal agents) use in the past 60 days for infection
29. History of an anaphylactic reaction or known sensitivity or intolerance to parenteral administration of contrast agents, human or murine proteins, or monoclonal antibodies, including rituximab or belimumab
30. Evidence of serious suicide risk including any history of suicidal behavior in the last 6 months and/or any suicidal ideation in the last 2 months, or who in the investigator's judgment, poses a significant suicide risk
31. Evidence of current drug or alcohol abuse or dependence, or a history of drug or alcohol abuse or dependence in the past 12 months
32. Vaccination with a live vaccine within the past 30 days
33. Other diseases or conditions or other clinically significant abnormal laboratory value which in the opinion of the investigator would put the patient at risk or confound the results of the study
34. Inability to comply with study and follow-up procedures

#### Section 2: Definitions for clinical outcomes

The definitions of complete and partial remission were updated for protocol v7.0 (July 29, 2024) to align with the definitions used in prior PMN studies, such as MENTOR.<sup>3</sup> The updated definitions and results are provided in the main manuscript. The original definitions along with corresponding results are provided here:

##### Original definition for complete remission (CR):

CR is defined as proteinuria of $\leq 0.3$ g/day with a $< 20\%$ decrease in estimated glomerular filtration rate (eGFR) from baseline.

##### Original definition for partial remission (PR):

PR is defined as a 50% or greater decrease in proteinuria compared to baseline and proteinuria $< 3.5$ g/day with a $< 20\%$ decrease in eGFR from baseline.

The clinical outcomes for Part A participants using the original definitions for CR and PR are presented in Supplemental Table S1. The clinical outcomes at week 104 are the same between the original and revised definitions of CR and PR.

Supplemental Table S1. Frequency of CR and PR at weeks 52 and 104 using the original definitions of CR and PR.

|  | Week 52 |  | Week 104 |  |
| --- | --- | --- | --- | --- |
| Study population | Treated<br>N=17 | Per Protocol<br>N=10 | Treated<br>N=17 | Per Protocol<br>N=11 |
| CR or PR, n (% achieved) | 5 (29%) | 5 (50%) | 10 (59%) | 10 (91%) |
| CR, n (% achieved) | 0 | 0 | 2 (12%) | 2 (18%) |

The following definitions of relapse, worsening proteinuria and worsening kidney function are provided for clarification and were unchanged during REBOOT Part A.

##### Relapse:

Relapse is defined as a return of proteinuria $\geq 3.5$ g/day after achieving a CR, or achieving and maintaining a PR for at least 12 weeks.

##### Worsening Proteinuria:

Worsening proteinuria is defined as an increase in proteinuria by $> 30\%$ from baseline with a corresponding decrease in serum albumin by $> 20\%$ from baseline.

##### Worsening Renal Function:

Worsening renal function is defined as a decrease in eGFR $> 25$ mL/min/1.73m<sup>2</sup> during the previous 52 weeks due to primary MN. The decrease in eGFR will be calculated using the maximum eGFR observed in the previous 52 weeks. The earliest time point for this assessment will be Visit 0. Other causes for declining renal function (e.g., concurrent acute illness, intravascular depletion, renal vein thrombosis, drug associated kidney injury from non-protocol medication) should be excluded. If a participant fulfills the definition of worsening renal function (including the confirmatory assessment), the site investigator should confer with the Protocol Chair to discuss whether the cause of the worsening renal function is due to primary MN.

##### Section 3: Additional information for flow cytometry assays

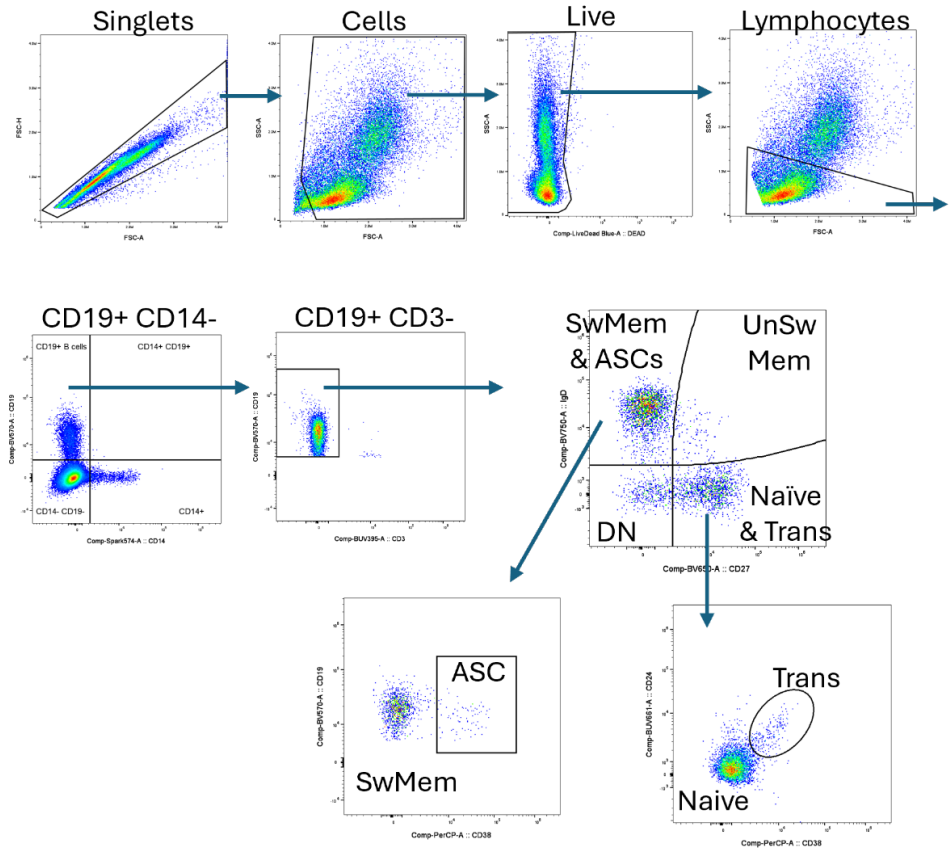

Supplemental Figure S1: Representative flow cytometric gating of B cell subsets, starting with clean up steps to remove doublets, dead cells, and non-lymphocytes, followed by gating on CD19+ CD14- CD3- B cells. Within B cells, CD27 and IgD were used to create four subsets: switched memory + antibody secreting cells (SwMem & ASCs), unswitched memory (UnSwMem), naïve & transitional (Naïve & Trans), and double negative (DN). Switched memory and ASCs were subdivided using CD38 expression, while naïve and transitional were subdivided using CD38 and CD24.

Supplemental Table S2: Antibody panel for B cell subset analysis by spectral flow cytometry

| Fluorochrome | Antigen | Clone |
| --- | --- | --- |
| BUV395 | CD3 | UCHT1 |
| BUV496 | CD8 | RPA-T8 |
| BUV563 | CD86 | 2331 (FUN-1) |
| BUV615 | CD95 | DX2 |
| BUV661 | CD24 | ML5 |
| BUV737 | CD127 | HIL-7R-M21 |
| BUV805 | CD56 | NCAM16.2 |
| BV421 | ICOS | C398.4A |
| BV480 | CD4 | RPA-T4 |
| BV510 | CD45RA | HI100 |
| BV570 | CD19 | HIB19 |
| BV605 | CD40 | 5C3 |
| BV650 | CD27 | O323 |
| BV711 | CD21 | B-ly5 |
| BV750 | IgD | IA6-2 |
| BV785 | PD-1 | EH12.2H7 |
| BB515 | CXCR5 | RF8B2 |
| RB545 | IgG | G18-145 |
| Spark Blue 574 | CD14 | HCD14 |
| BB630-P | FOXP3 | 259D/C7 |
| BB660-P | Ki67 | B56 |
| PerCP | CD38 | HIT2 |
| BB700 | CCR7 | 3D12 |
| PerCP-eF710 | IgM | SA-DA4 |
| RB744 | FCRL5 | 509F6 |
| RB780 | BAFF-R | 11C1 |
| PE | TACI | 1A1 |
| RY610 | HLA-DR | L203 |
| PE-Cy5 | CD25 | M-A251 |
| PE-Cy7 | BCMA | 19F2 |
| AF647 | CD138 | MI15 |
| AF700 | CD11c | Bu15 |
| APC-eFluor780 | IgA | HM47 |
| APC/FIRE 810 | CD20 | S18015E |

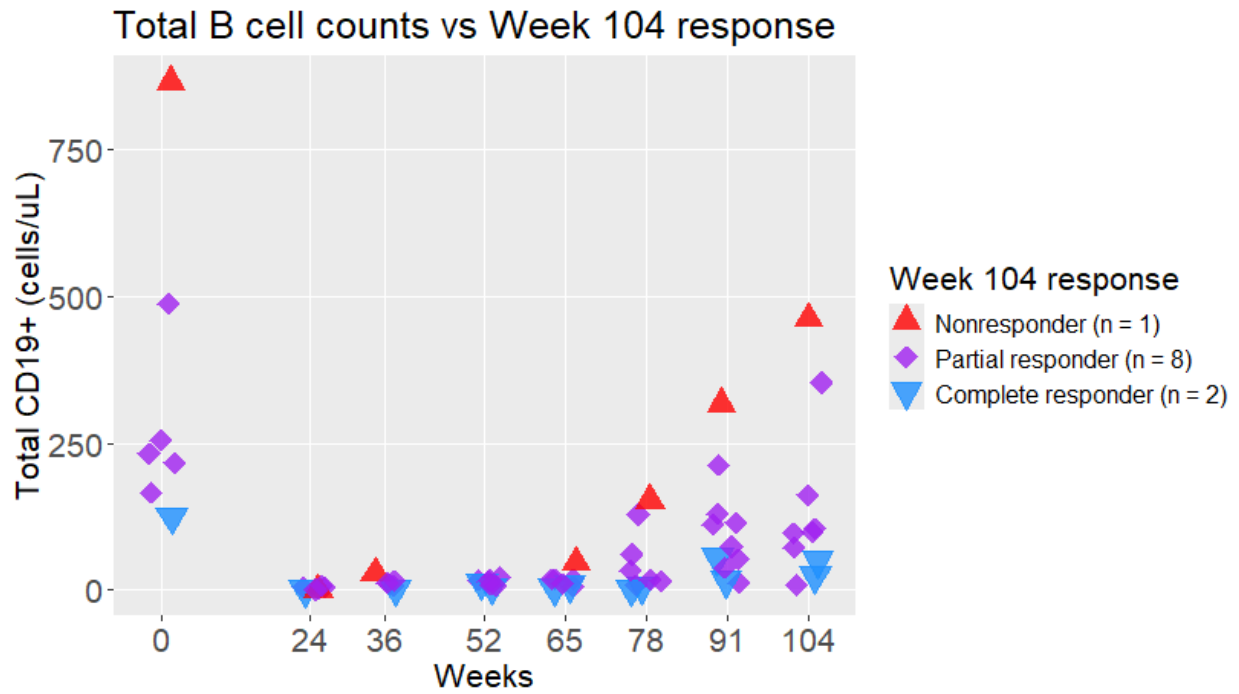

**Supplemental Figure S2.** Total CD19+ B cell counts for participants based on response status from enrollment to week 104.

###### Section 4: Participant discontinuation

Participants discontinued BEL if they did not receive at least one dose of RTX, developed worsening kidney function due to PMN, had worsening proteinuria, experienced a relapse, used a prohibited medication, experienced a grade 3 infection in the setting of grade 4 hypogammaglobulinemia, became pregnant, if the participant requested to halt treatment, or if the investigator determined that it was in the participant's best interest.

Five participants did not complete belimumab (BEL) dosing and entered safety monitoring. The details regarding why they did not complete BEL dosing are provided below:

1. One participant was discontinued after week 1 due to sponsor decision. This participant enrolled just prior to the start of the COVID-19 pandemic. Given potential risks of infection with immunosuppression as well as lack of therapies available to treat COVID-19, the sponsor elected to discontinue BEL. The participant did not receive rituximab.
2. One participant discontinued BEL due to a grade 3 infection in the setting of Grade 4 hypogammaglobulinemia. The participant developed Grade 3 infective bursitis in the setting of Grade 4 hypogammaglobulinemia (serum IgG 245 mg/dL at baseline). BEL was discontinued after week 2, and the participant did not receive rituximab.
3. One participant discontinued BEL after week 7 at the investigator's discretion. The participant developed significant mood swings while on BEL, and the investigator felt it was in the participant's best interest to discontinue BEL.
4. One participant discontinued from BEL after week 9 due to worsening renal function. The estimated GFR for this participant during study participation is shown in Supplemental Table S3.

Supplemental Table S3. eGFR for participants who discontinued due to worsening renal function.

| Study Visit | Estimated GFR<br>(mL/min/1.73m<sup>2</sup>) |
| --- | --- |
| Screening | 56.9 |
| Day 0 | 34.8 |
| Week 4 | 32.6 |
| Week 6 | 18.8 |
| Week 8 | 27.3 |

5. One participant discontinued BEL due to too many consecutive missed doses due to a COVID-19 infection. The participant missed 4 consecutive doses of BEL and was formally discontinued from BEL after week 32.

**Section 5:** Treatment-Emergent adverse events with Incidence $\geq 10\%$ of participants:

Supplemental Table S4. Treatment-Emergent AEs with Incidence $\geq 10\%$.

| Body System<br>Preferred Term [1] | Participants<br>(N=17) | Incidence (%) |
| --- | --- | --- |
| Investigations |  |  |
| Glomerular filtration rate decreased | 6 | 35 |
| Blood creatinine increased | 4 | 24 |
| Neutrophil count decreased | 3 | 18 |
| Immune system disorders |  |  |
| Hypogammaglobulinaemia | 8 | 47 |
| Infections and infestations |  |  |
| COVID-19 | 6 | 35 |
| Upper respiratory tract infection | 2 | 12 |
| Blood and lymphatic system disorders |  |  |
| Neutropenia | 3 | 18 |
| Metabolism and nutrition disorders |  |  |
| Hyponatraemia | 2 | 12 |
| Psychiatric disorders |  |  |
| Abnormal dreams | 2 | 12 |
| Vascular disorders |  |  |
| Hypertension | 2 | 12 |

[1] MedDRA Version 23.0

### The STROBE reporting checklist

For checking that observational epidemiology research articles can be understood and used by everyone

#### Note

If you have not used a reporting guideline before, read about how and why to use them and check whether STROBE is the most applicable reporting guideline for your work.

Reporting guidelines are most useful when used early in research. When writing a manuscript or application, consider using the Full Guidance where you'll see explanations and examples for each item.

After writing, demonstrate adherence by completing this checklist:

1. Specify where each item is described (see Note 1).
2. Cite this checklist (See Note 2).
3. Include your completed checklist as a supplement when submitting to a journal so that future readers can use it to find information.

|  | Item Description | Location (or reason for not reporting) |
| --- | --- | --- |
| **Title and abstract** |  |  |
| 1a. Indicate the study's design | Indicate the study's design with a commonly used term in the title or the abstract. | Abstract |
| 1b. Abstract | Provide in the abstract an informative and balanced summary of what was done and what was found. | Abstract |
| **Introduction** |  |  |
| 2. Background / rationale | Explain the scientific background and rationale for the investigation being reported. | Introduction: paragraphs 1-3 |
| 3. Objectives | State specific objectives, including any prespecified hypotheses. | Introduction: paragraph 4, Methods: paragraph 1 |
| **Methods** |  |  |
| 4. Study design | Present key elements of study design early in the paper. | Methods: Study participants, Study treatment and clinical assessments |
| 5. Setting | Describe the setting, locations, and relevant dates, including periods of recruitment, exposure, follow-up, and data collection. | Methods: Study participants: paragraph 1, Study treatment and clinical assessments; Results: student enrollment paragraph 1 |
| 6a. Eligibility criteria | **Cohort study:** Give the eligibility criteria, and the sources and methods of selection of participants. Describe methods of follow-up. **Case-control study:** Give the eligibility criteria, and the sources and methods of case ascertainment and control selection. Give the rationale for the choice of cases and controls. **Cross-sectional study:** Give the eligibility criteria, and the sources and methods of selection of participants. | Methods: Study participants; Supplemental Materials: Section 1 |
| 6b. Matching criteria | **Cohort study:** For matched studies, give matching criteria and number of exposed and unexposed. **Case-control study:** For matched studies, give matching criteria and the number of controls per case. | Not applicable. |
| 7. Variables | Clearly define all outcomes, exposures, predictors, potential confounders, and effect modifiers. Give diagnostic criteria, if applicable. | Methods: Study treatment and clinical assessments |
| 8. Data sources / measurement | For each variable of interest give sources of data and details of methods of assessment (measurement). Describe comparability of assessment methods if there is more than one group. | Methods: Study treatment and clinical assessments |
| 9. Bias | Describe any efforts to address potential sources of bias. | Discussion: paragraph 6 |
| 10. Study size | Explain how the study size was arrived at. | Methods: statistical analysis |
| 11. Quantitative variables | Explain how quantitative variables were handled in the analyses. If applicable, describe which groupings were chosen, and why. | Methods: statistical analysis |
| 12a. Statistical methods | Describe all statistical methods, including those used to control for confounding. | Methods: statistical analysis |
| 12b. Statistical methods – subgroups and interactions | Describe any methods used to examine subgroups and interactions. | Not applicable. |
| 12c. Statistical methods – missing data | Explain how missing data were addressed. | Methods: statistical analysis (clinical and patient reported outcomes) |
| 12di. Statistical methods – loss to follow-up | **Cohort study:** If applicable, describe how loss to follow-up was addressed. | Methods: statistical analysis (clinical and patient reported outcomes) |
| 12dii. Statistical methods – matching cases and controls | **Case-control study:** If applicable, explain how matching of cases and controls was addressed. | Not applicable |
| 12diii. Statistical methods – sampling strategy | **Cross-sectional study:** If applicable, describe analytical methods taking account of sampling strategy. | Not applicable |
| 12e. Statistical methods – sensitivity analyses | Describe any sensitivity analyses. | Not applicable |
| **Results** |  |  |
| 13a. Participant numbers | Report the numbers of individuals at each stage of the study—e.g., numbers potentially eligible, examined for eligibility, confirmed eligible, included in the study, completing follow-up, and analysed; Consider use of a flow diagram. | Figure 1 |
| 13b. Participants – non-participation | Give reasons for non-participation at each stage. | Figure 1; Supplemental material: Section 4 (participant discontinuation) |
| 13c. Participants – flow diagram | Consider use of a flow diagram. | Figure 1. |
| 14a. Descriptive data – participant characteristics | Give characteristics of study participants (e.g., demographic, clinical, social) and information on exposures and potential confounders. Present the information in a table. | Table 1. |
| 14b. Descriptive data – missing data | Indicate the number of participants with missing data for each variable of interest. | Table 2; Results: clinical and patient reported outcomes |
| 14c. Descriptive data – follow-up time | **Cohort study:** Summarise follow-up time—e.g., average and total amount. | Results: clinical and patient reported outcomes |
| 15. Outcome data | **Cohort study:** Report numbers of outcome events or summary measures over time. **Case-control study:** Report numbers in each exposure category, or summary measures of exposure. **Cross-sectional study:** Report numbers of outcome events or summary measures. | Table 2, Table 3; Results: Belimumab exposure analysis, clinical and patient reported outcomes |
| 16a. Main results | Give unadjusted estimates and, if applicable, confounder-adjusted estimates and their precision (e.g., 95% confidence intervals). Make clear which confounders were adjusted for and why they were included. | Table 2, Table 3; Results: Belimumab exposure analysis, clinical and patient reported outcomes |
| 16b. Main results – category boundaries | Report category boundaries when continuous variables were categorised. | Not applicable. |
| 16c. Main results – risk | If relevant, consider translating estimates of relative risk into absolute risk for a meaningful time period. | Not applicable. |
| 17. Other analyses | Report other analyses done—e.g., analyses of subgroups and interactions, and sensitivity analyses. | Not applicable. |
| **Discussion** |  |  |
| 18. Key results | Summarise key results with reference to study objectives. | Discussion: paragraph 1 and 2 |
| 19. Limitations | Discuss limitations of the study, taking into account sources of potential bias or imprecision. Discuss both direction and magnitude of any potential bias. | Discussion: paragraph 6 |
| 20. Interpretation | Give a cautious overall interpretation considering objectives, limitations, multiplicity of analyses, results from similar studies, and other relevant evidence. | Discussion: paragraphs 4 and 6 |
| 21. Generalisability | Discuss the generalisability (external validity) of the study results. | Discussion: paragraph 6 |
| **Other information** |  |  |
| 22. Funding | Give the source of funding and the role of the funders for the present study and, if applicable, for the original study on which the present article is based. | Funding (page 1). |
